# Using Machine Learning along with Data Science algorithms to pre-process and forecast COVID-19 Cases and Deaths

**DOI:** 10.1101/2021.03.15.21253571

**Authors:** Avi Choudhary

## Abstract

The Covid-19 pandemic has taken a major toll on the health and state of our global population. With tough decisions for allocating resources(i.e. vaccines)[1] are being made, forecasting through machine learning has become more important than ever. Moreover, as vaccines are being brought to the public and cases are going down, it is time that we reflect on where the pandemic has taken the most toll:for the purpose of future reform. This research illustrates two different models and algorithms for COVID-19 forecasting: Auto Regressive models and Recurrent Neural Networks(RNNs). The results show the true potential of RNNs to work with sequential and time-series data to forecast future cases and deaths in different states. As the paper utilizes the tanh activation function and multiple LSTM layers, the research will show the importance of machine learning and its ability to help politicians make decisions when it comes to helping states during the pandemic and future reform. The data will also pre-process the time-series data, using rolling statistics and will clean the data for the auto-regressive model and RNN layers. Thus, we show that along with Recurrent Neural Network layers, activation functions also play a crucial role in the accuracy of the forecast.

## Introduction

The pandemic has taken its toll on the global population, taking the lives of many loved ones, while placing many people in poverty. With the United States having one of the largest populations in the world, the virus displayed its influence drastically in the country. Around 30 million people suffered at the hands of the virus, and over 500,000 people have died from the disease[2]. As this trend was previously increasing at a drastic rate, prediction and forecasting models are as important and crucial to our society as ever. The public needs to understand which states have suffered the most at the hands of the virus in order to prevent this catastrophe in the future[3]. Through forecasting the cases and deaths, it is extremely crucial that we are not able to just forecast the future cases and deaths, but are able to do so accurately.

As auto-regressive models and deep learning Recurrent Neural networks can help us in predicting future cases and deaths, its accuracy will require an activation function of somesort. Thus, it is crucial that we use activation functions to help the model adjust greatly towards the time-series data. A medRxiv paper shows using Recurrent Neural Networks using the sigmoid activation function; however, their research can also be expanded by using the tanh function, which is better used for Recurrent Neural Networks[4]. Given the vast amount of COVID-19 data provided, data collection is not the problem. However, the COVID-19 data does have a lot of null values and inaccurate data, so pre-processing the data is also crucial. Furthermore, the auto-regressive model needs to have data that is stationary for the model to produce better results, thus weighted moving averages will be needed for the model.

The goal of this research is to illustrate how vital activation functions and forecast algorithms are important in our society. As the data will be thoroughly pre-processed and applied to the network, the results will show how the virus has exponentially increased the number of cases and deaths.

## Methods

This research focuses on forecasting future COVID-19 data using auto-regressive models and Recurrent Neural Networks: using a Kaggle dataset from the New York Times that initially had 6 columns—date, county, state, fips, cases, deaths— with around 100,000+ rows[5]. The data reaches till December 2020, so we can predict that the cases and deaths should still be increasing. To clean the data, the county and fips column had to be dropped. The dataset was then combined by state, rather than county. Furthermore, the null values were dropped and the final results were graphed.

**Figure1:**
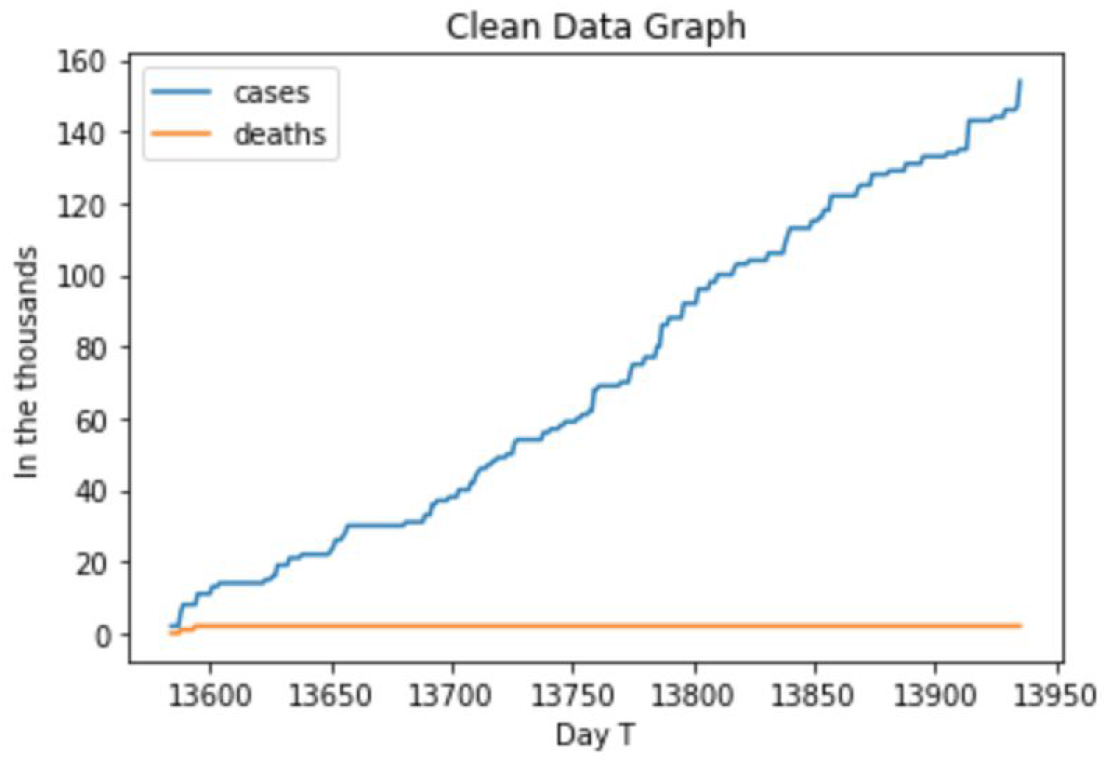
Visualization of the clean data’s cases and deaths.

For the auto-regressive model, the data had to be made stationary to produce more accurate results. To make sure that the data was stationary, the research implemented rolling statistics which used the rolling mean to make the data more stationary. We used the weighted moving average to give unique weights to different data points.

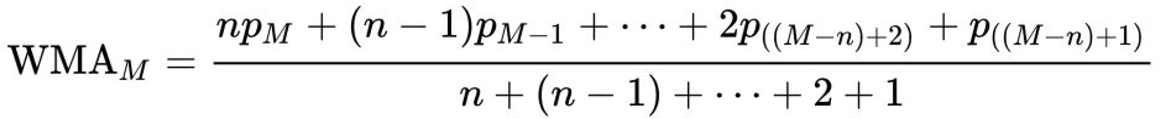

The numerator represents the points being multiplied by certain weights(increasing as the time/points goes by). The research had used the weighted moving average, as we wanted to place more of an importance on the new data points rather than the old data points. This will clearly tell the auto-regressive model what the trend clearly is as this will smooth the data more.

As we are taking advantage of the time-series data, our goal is to predict the future cases and deaths i + 7th days ahead. Implementing the auto-regressive model is fairly simple, however, adjusting to the shape of the data and shifting the data is quite intricate. As the data requires manipulation of the data shape to fit in the model, the process can get extremely intricate and complex. The equation for the auto-regressive model is as follows:

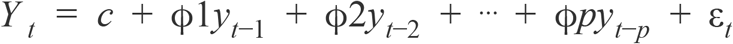

Later, Recurrent Neural Network layers were applied to the pre-processed data and the LSTM layers were set to the current hyperparameters. The LSTM layers are essential as they use the past sequence memory needed to predict the future sequence. The mathematical equations in the LSTM network is:

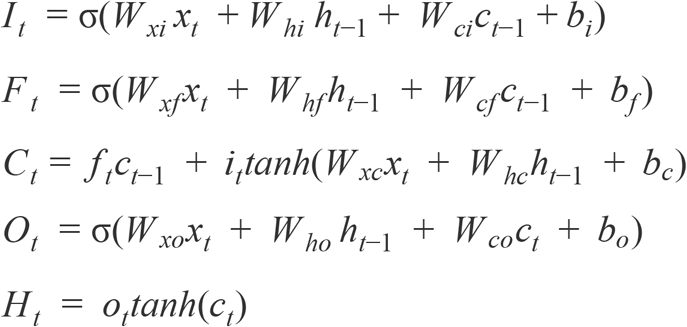

The *i* is the input gate; the *f* represents the forget gate; the *c* is the memory cell, and the *o* is the output gate. As implementing multiple LSTM layers would be too convoluted a single LSTM layer was used. We used the stacked LSTM layer, whiched called for two LSTM layers and one dense layer. We used two activation functions: the sigmoid(σ) and the tanh function in order to see how the RNN layers react to the functions.

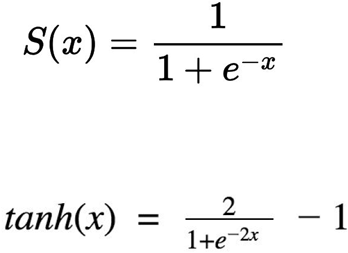

The train and test data split was 80-20, which allowed us sufficient data for us to train the model. The 20% of the dataset would be used to test how well the model did with new data. We can check how well the data adjusts to the model through a loss function. Below is the equation of the cross entropy loss function implemented.

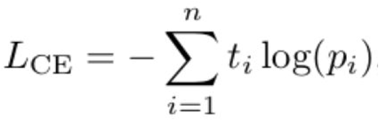

## Results

As we mentioned before, the model’s error can be measured through different loss functions. In our research, we used the Root Mean Square error to determine the loss.

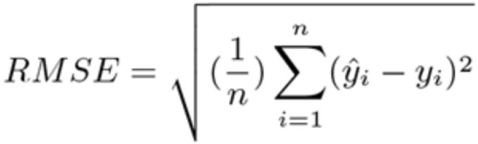

For this paper, we will only include one state: California. The RMSE for the auto-regressive model in forecasting the future US COVID-19 deaths is 999.1466 and the cases is 7629.379. The RMSE for the recurrent neural networks implementation, along with the activation function for the deaths is 612.3913 and the cases is 5491.7842. The auto-regressive value predictor is 0.765 and the predictor for the recurrent-neural network model is 0.883. The figure below shows the cases and deaths prediction for the state of California using the Recurrent Neural Network model, along with its activation functions.

**Figure 2:**
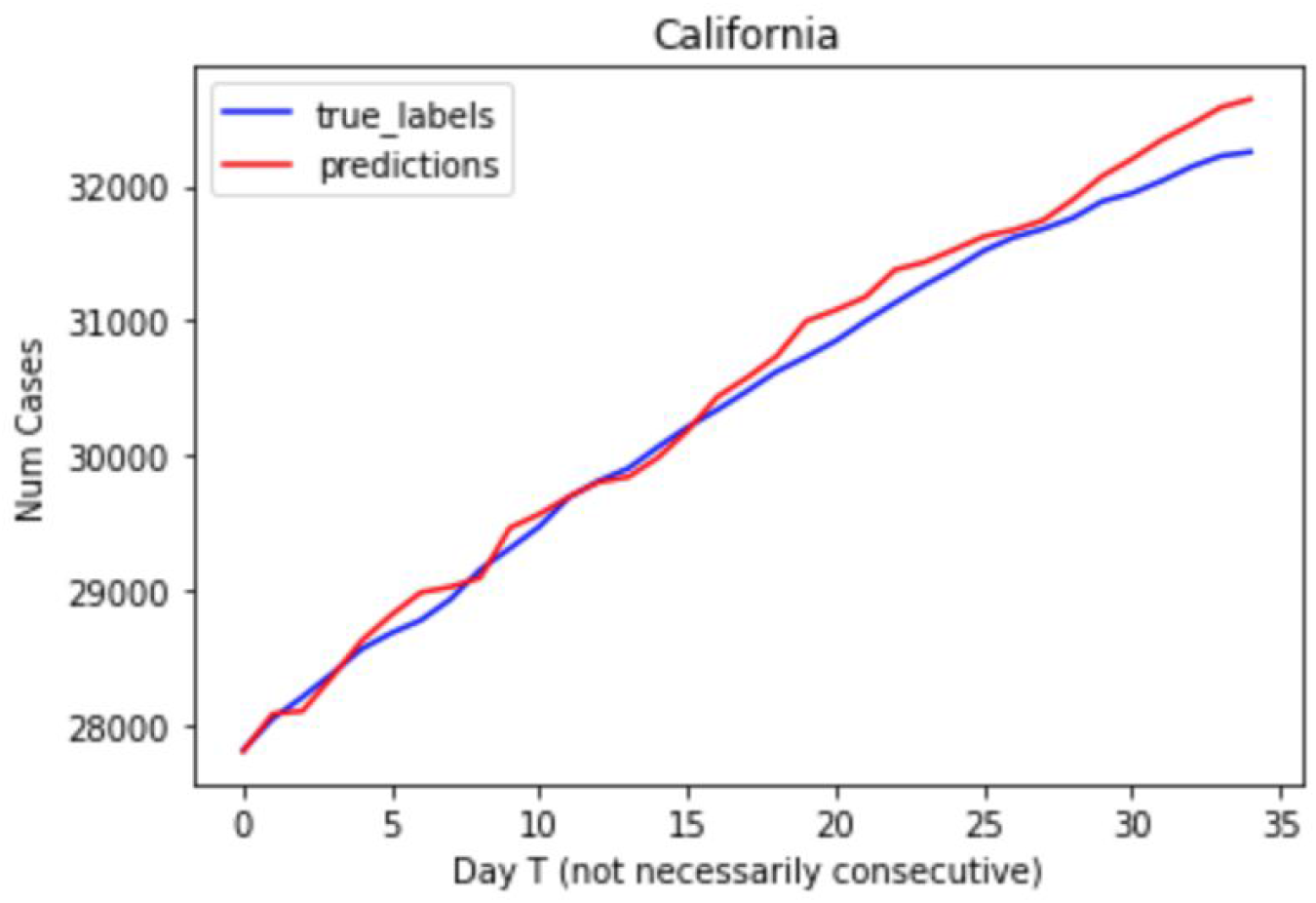
Here the cases’ true labels and the RNNs model’s predictions are graphed.

**Figure 3:**
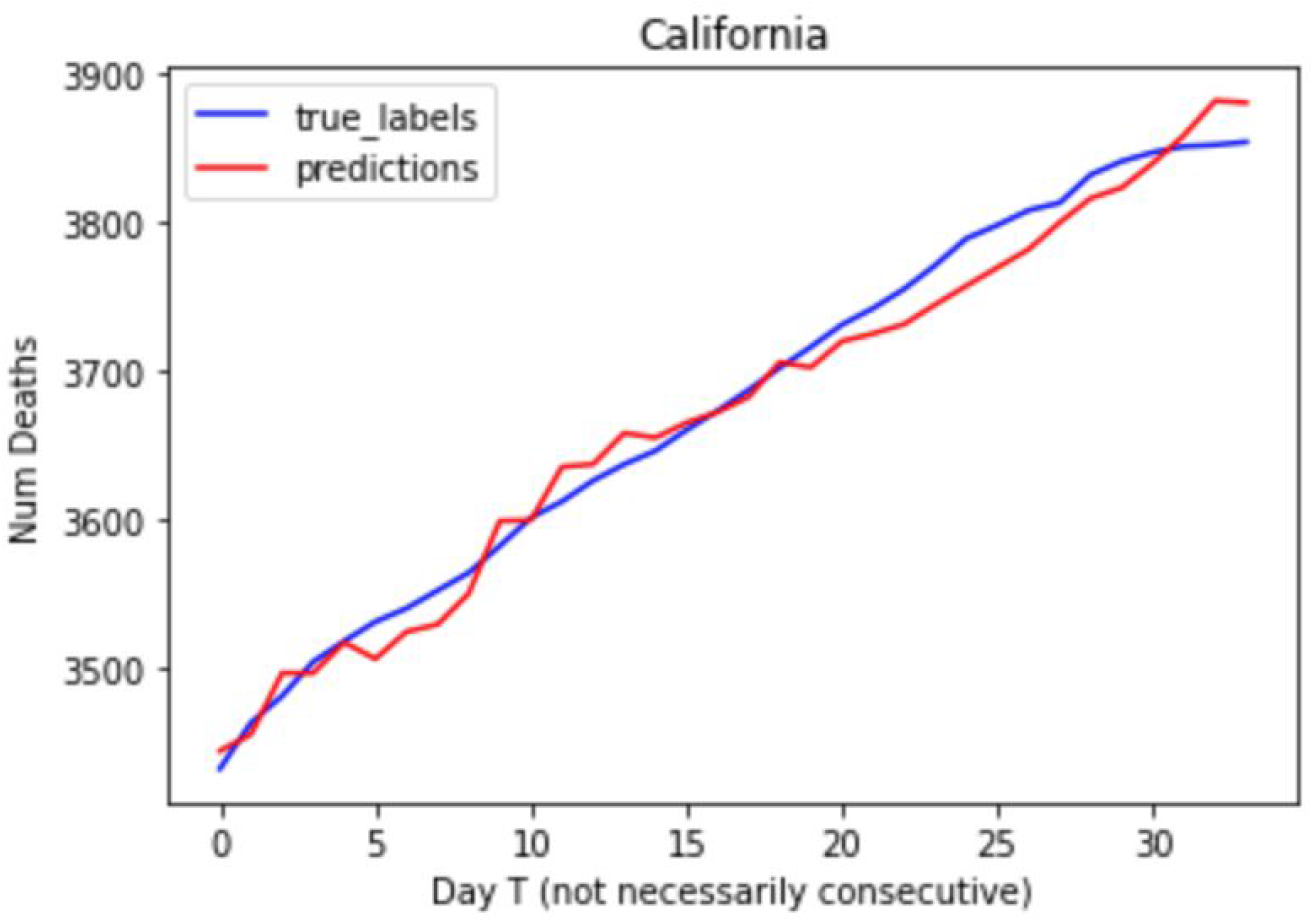
A visualization that represents the number of deaths’ true labels vs the RNN model’s predictions.

## Discussion

The figures clearly represent the accurate predictions that the RNNs forecast regarding the future COVID-19 Cases and Deaths.

We can see that compared to the auto-regressive model, the RNNs have a much better RMSE error as the deep learning algorithm uses Long Short Term layers and activation functions for the model to better adjust to the time-series data. The research did not just implement a sigmoid function, but also imported a tanh function to better help the RNN model adjust to the data. Furthermore, adding the weights to average out the data significantly helped to adjust to the model. While the data did not need to be averaged for the RNNs, as it has the ability to adjust to the data, it was well needed for the auto-regressive model.

While there is still improvement needing to be made in the RNNs implementation, as the forecast accuracy should be around > 0.995 for the model to be beneficial, the results still show that the model is more useful and beneficial towards forecasting rather than the auto-regressive model. We can also see that the RMSE error differs between the cases and deaths as the deaths error is lower. However, the number of deaths have different training data and numbers compared to the number of deaths, so we cannot compare the two RMSE errors. The dataset was till December 2020, and so we can still see that the cases and deaths are rising and the model as well shows the similar trend.

Even looking at a simple average model from other papers, we can see that the RNN application has produced much better results, indicating that neural networks produce the depth needed to produce accurate results[6].

As the graphs show an increase in the cases and deaths in California that reach statistically high numbers, the government should take into account that because California has a large population, they should have the right to more resources and vaccines. Even in the future, better precautions should be taken to prevent California from becoming one of the deadliest states in the country.

## Conclusion

The US has shown the world how easily it is susceptible to the virus, and how deadly the virus is. This paper focuses on the huge benefits of using deep learning algorithms and activation functions rather than simple auto-regressiv models. While this research does focus on auto-regressive models, the dataset can also be applied to Auto-regressive integrated moving average models as well; we can check what the difference in the results are when compared to the RNNs. There is still improvement for the RNN models as the layers’ parameters could be better classified and implemented. We can also use convolutional neural networks to better improve the accuracy of the model by using 2D convolutional layers. Furthermore, we can go even more in depth and explore by county. By doing this, the public will know which area of the state is contributing to the spread of the virus(i.e Los Angeles county in California).

## Data Availability

The availability of all the data referred to in the manuscript and note links below.

https://www.kaggle.com/fireballbyedimyrnmom/us-counties-covid-19-dataset

## Data Availability

The raw COVID-19 Time-series data, created by the New York Times, is currently available in the Kaggle repository.

https://www.kaggle.com/fireballbyedimyrnmom/us-counties-covid-19-dataset

